# Comprehensive Benchmarking of CNN-Based Tumor Segmentation Methods Using Multimodal MRI Data

**DOI:** 10.1101/2024.01.22.24301602

**Authors:** Kavita Kundal, K Venkateswara Rao, Arunabha Majumdar, Neeraj Kumar, Rahul Kumar

## Abstract

Magnetic resonance imaging (MRI) is become an essential and a frontline technique in the detection of brain tumor. However, manual segmentation of tumor from MRI scans is a time-consuming and labour-intensive process. There is a prevalent trend in employing fully automated methods for accurate tumor segmentation using MRI scans. The precision in brain tumor segmentation is essential for the better diagnosis, treatment and prognosis. This study focuses on benchmarking and evaluating the performance of four widely used convolutional neural network (CNN) based brain tumor segmentation methods CaPTk, 2DVNet, EnsembleUNets, and ResNet50. We used 1251 multimodal MRI scans from the BraTS2021 dataset, which encompasses T1, T2, T1ce, and Flair imaging modalities. We compared the performance of these CNN-based methods against a reference set of previously segmented images obtained with the assistance of radiologists. The evaluation encompasses direct utilization of segmented outputs and also by employing radiomic features. Performance evaluation with direct segments method using Dice Similarity Coefficient score (DSC) and Hausdorff Distance (HD) suggested better performance of EnsembleUNets with DSC and HD of 0.93 and 18 respectively, outperforming the other methods. Comparative analysis using radiomics features also revealed that EnsembleUNets is the most precise segmentation method as compared to CaPTk, 2DVNet, and ResNet50. EnsembleUNets achieved Concordance Correlation Coefficient (CCC), Total Deviation Index (TDI), and Root Mean Square Error (RMSE) of 0.79, 1.14 and 0.53 respectively and outperformed its counterparts. These findings contribute valuable insight into the comparative efficacy of EnsembleUNets, facilitating informed decision for accurate brain tumor segmentation.

## 1. Introduction

Magnetic Resonance Imaging (MRI) has emerged as a cornerstone in the diagnosis and treatment planning of various medical conditions, particularly in the field of neuro-oncology (Sindhu et al., 2022). In the context of gliomas, which constitute approximately 80% of malignant brain tumors, the importance of MRI cannot be emphasized enough. Gliomas originate from crucial brain tissues, including oligodendrocytes, ependymal cells, and astrocytes, necessitating a comprehensive understanding of their molecular and pathological characteristics for precise diagnosis and treatment. The classification of gliomas into different grades, ranging from I to IV, reflects their diverse histological appearances and malignant potential (Louis et al., 2021). Transition from proneural to mesenchymal subtype in glioblastoma patients has been suggested as a key mechanism of tumor resistance to treatment (Duhamel et al., 2022). Patients diagnosed with mesenchymal subtype have shown worse survival outcomes than other subtypes (Z. Zhang et al., 2021). Molecular characteristics of the tumor help determine whether the patient will respond to treatment. Therefore, knowledge about molecular level status is essential for precision treatment and predicting the survival outcome.

The molecular characterization and pathological confirmation of brain tissue affected by tumors through invasive methods such as biopsy or surgical resection are not practically feasible (Puig et al., 2023). In contemporary medical practices, non-invasive techniques, particularly radiological imaging like magnetic resonance imaging (MRI), have become the most prevalent means of assessing the entire brain for diagnostic and treatment planning purposes (Shui et al., 2021). The evolution of radiomics has significantly contributed to transforming radiological images into analysable data, enabling the extraction of diagnostic and prognostic information (Habib et al., 2021; Singh et al., 2021). In the current research, investigators employ multimodal MRI images to depict glioblastoma’s intrinsic heterogeneity and phenotype (Cui et al., n.d.; Ye et al., 2021). Regions exhibiting hypo/hyper-intensity in various MRI modalities are crucial in providing complementary profiles of glioblastoma subregions (Ellingson, 2015; Liu et al., 2021). Extracting radiomic features from the tumor portion is imperative to obtain clinically relevant information for early diagnosis and treatment planning. Achieving accurate tumor segmentation from MRI scans is critical in this process. The precision of tumor segmentation directly influences the potency of the features derived from it (Ranjbarzadeh et al., 2021). Hence, the selection of the optimal method for tumor segmentation holds paramount importance.

The conventional brain tumor segmentation method from pre-operative MRI scans, performed by neuroradiologists, is often challenged by high variability in shape and appearance, ambiguous boundaries, and imaging artifacts, leading to a time-consuming and challenging process (Bhandari et al., 2020). Addressing this,“Automatic tumor segmentation”utilizing deep learning methods has gained significant success in medicine. Among these methods, Convolutional Neural Networks (CNNs) with multiple hidden convolutional layers and activation layers have demonstrated excellent results in various oncogenic detection tasks, including skin cancer classification (Fu’adah et al., 2020), brain tumor segmentation (Sun et al., 2019), NSCLC classification(Chen et al., 2021), and breast cancer detection(Desai & Shah, 2021). Despite the success, reliability, and accessibility of different methods for performing brain tumor segmentation using deep learning methods have not been systematically benchmarked. In this current study, we address the existing gap by providing a comprehensive benchmarking analysis that compares the accuracy and robustness of widely used brain tumor segmentation methods.This study aims to systematically evaluate the reliability and accessibility of four different widely used automated deep learning methods for brain tumor segmentation i.e., CaPTk software (Rathore et al., 2018), 2DVNet (Rastogi et al., 2021), Ensemble UNets (Y. Zhang et al., 2021), and ResNet50 (Thhntb. (2021), n.d.). The proposed framework encompasses three key steps: tumor segmentation, radiomic feature extraction, and performance analysis as shown in **Figure 1**. By rigorously evaluating these methods, this research aims to identify the most accurate and robust automated method for brain tumor segmentation. These methods enhance early detection, facilitates treatment planning, and ultimately improves patient survival outcomes by providing accurate insights for neurosurgeons and oncologists. The efficiency and consistency of automated segmentation also contribute to reducing diagnostic errors and streamlining the healthcare workflow.

**Figure 1:**
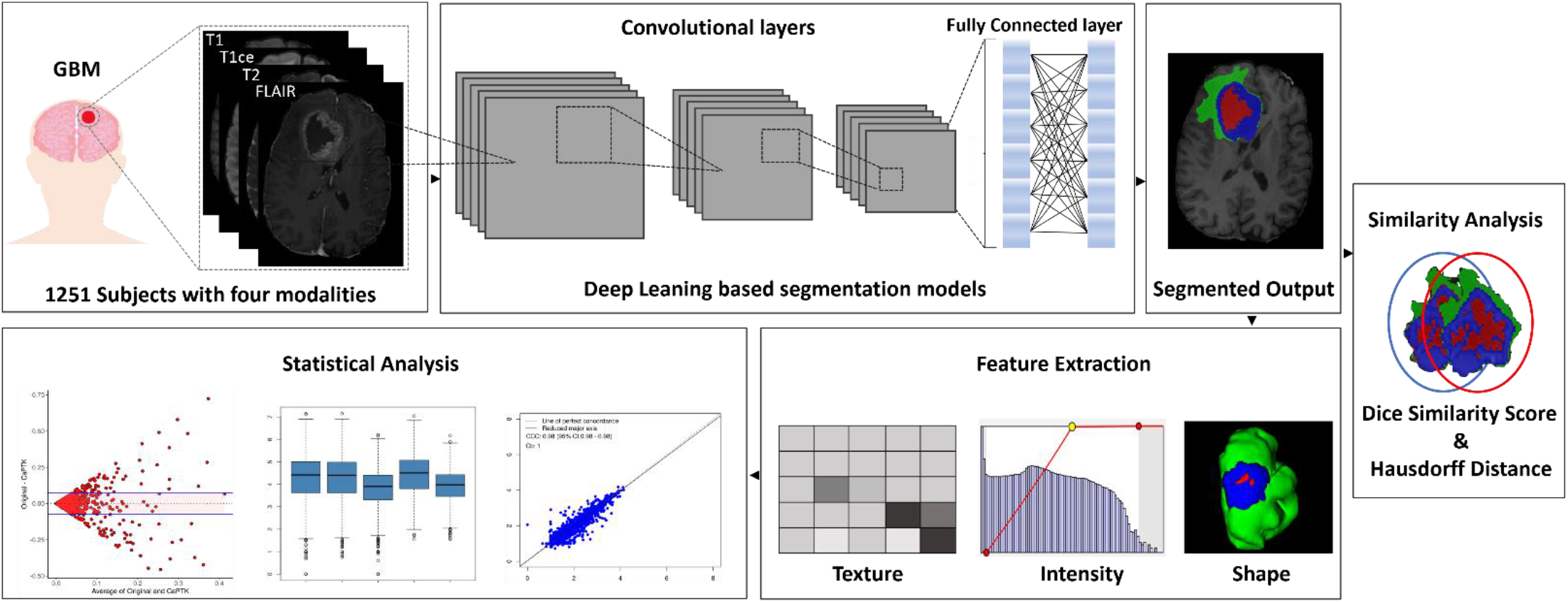
Overview of the study illustrating data description, deep learning segmentation, feature extraction, and statistical evaluation for performance assessment.

## 2. Methodology

### 2.1 Data description

The dataset used in this study has been downloaded from the RSNA-ASNR-MICCAI Brain Tumor Segmentation Challenge 2021 (*https://Www.Rsna.Org/Rsnai/Ai-Image-Challenge/Brain-Tumor-Ai-Challenge-2021*, n.d.). BraTS2021 dataset comprises a total of 2000 subjects with 8000 mpMRI scans sourced from different institutions and studies such as TCGA-GBM, TCGA-LGG, Ivy-GAP, CPTAC-GBM, ACRIN-FMISO-Brain and some contributions from private institutional collections. Of these 2000 subjects, only 1251 subjects that consist of glioblastoma (GBM) and low-grade glioma (LGG) samples are available and were taken further for this study. MRI scans of all subjects have 4 different modalities, i.e., T1, T2, T1-contrast enhanced (T1ce), and FLAIR in NIfTI (.nii.gz) format with the dimension of 240×240×155. This study utilized pre-segmented scans from the BraTS2021 dataset as a reference for comparative analysis. The reference segmentation was executed using the STAPLE method which involved the integration of three advanced deep learning methods: nnUNET, DeepScan, and DeepMedic. To ensure the precision of the reference segmentation, it was validated by a radiologist. As shown in **Figure 2**, reference segmentation annotations consist of three subtypes i.e. Necrotic/ Non-enhancing tumor (NCR), Enhancing tumor (ET), and peritumoral edema (ED) (Baid et al., 2021; Bakas et al., 2017; Menze et al., 2015). These sub-regions aid in precise treatment planning by identifying specific areas of tumor infiltration and assists in studying the heterogeneous nature of glioma, leading to more targeted therapies and improved understanding of the disease progression.

**Figure 2:**
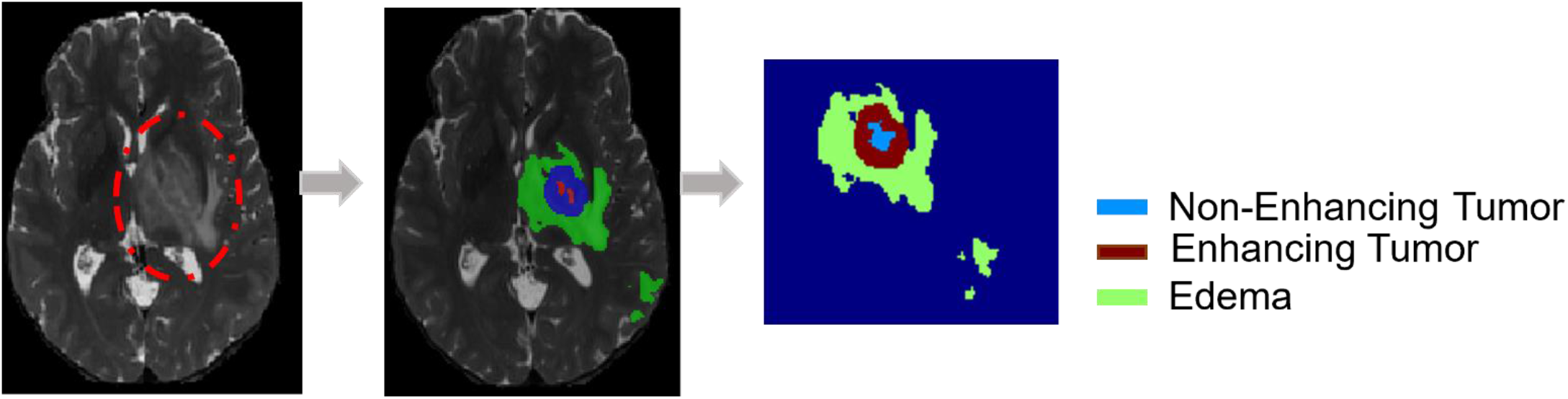
Glioma subregions visualization on MRI. A unified tumor segment is color-coded to showcase key subregions: Blue for necrotic regions (NET), red for enhancing tumor (ET), and green for peritumoral edema. This integrated representation enhances the clarity of glioma heterogeneity on MRI scans.

### 2.2 CNN based Segmentation Methods

We have adopted four well-known, widely used CNN-based segmentation methods to benchmarks using BraTS2021 dataset.

#### 2.2.1 CaPTk

The CNN method“DeepMedic”is pre-trained on BraTS2017 training data and is working at the backend of the CaPTk Brain Tumor Segmentation module. DeepMedic is an 11-layer brain lesion segmentation method based on a multi-scale 3D deep CNN coupled with a fully connected 3D conditional random field (CRF) (Kamnitsas et al., 2017). Each layer within the illustrated pathways consists of two layers employing 3^3^ kernels, exemplifying a comprehensive approach to hierarchical feature extraction in 3D data. The multi-scale 3D deep CNN comprises two convolutional pathways utilizing 53 kernels, which were later substituted with 3^3^ kernels in the DeepMedic architecture. Both pathways handle inputs centered at identical locations, while the second pathway operates on a down-sampled version, reduced by a factor of 3. Despite down-sampling, the second pathway considers a larger spatial context of 51^3^ voxels. The neurons in the final layers of both pathways exhibit receptive fields of 17^3^ voxels (Rathore et al., 2018). For this study we have used command-line with default parameters provided by CaPTk developers.

#### 2.2.2 2DVNet

Volumetric convolutional network (V-NET) is designed for medical image segmentation to address challenges in tumor segmentation from multimodal brain MRI images. The architecture of VNET consists of an encoder (left-side network) and decoder (right-side network) used for signal compression and decompression, respectively. It employs capturing spatial relationships and hierarchical features. Horizontal connections facilitate transferring features from encoder to decoder; this provides accurate location information and enhances the method’s convergence time. The Parametric Rectified Linear Unit (PReLU) is the activation function, allowing the network to learn more complex relationships and patterns in the data, also known as activating non-linearity. Downsampling is often used to broaden the receptive field. We used the pre-build 2D-Vnet method trained on the HGG BraTS2020 dataset using python3 and the output used for comparative analysis (Rastogi et al., 2021).

#### 2.2.3 EnsembleUNets

Unet is a convolutional neural network widely used in semantic segmentation tasks, including medical image segmentation. It has been proven the most effective method for brain tumor segmentation from multimodal MRI images. Unet model consists of a contracting path that captures context and helps reduce the spatial resolution; bottleneck helps in local and global feature extraction, context encoding, and information fusion. In contrast, the third component of Unet, i.e., expansive path, combines background information to generate a high-resolution segmentation map. These characteristics of UNets make it suitable for precise brain tumor segmentation from mpMRI images. Zhang et al. created an ensemble model by combining three independent models: 3D Unet, 3D MI-Unet, and 3D+2D MI-Unet (Wu et al., 2019). The input of the first model uses 3D patches of MRI images, the second model uses brain parcellation (BP), and the third model uses 2D slices and output from the second model. The first two models produce TC and ET segments fused with WT segmentation. Jointly working all three ensembled UNets in fully automated ways improves segmentation accuracy and works most effectively after adopting a post-processing strategy to label ET as a non-enhancing tumor to give correct segmentations. We implemented the pre-trained model (BraTS2018 dataset) submitted in docker container *“sustechmedical/brain_tumor_segmentation”* using Docker pull and run command to get tumor segments directly from pre-processed input data (Y. Zhang et al., 2021).

#### 2.2.4 ResNet50

ResNet is a semantic segmentation model developed using TensorFlow/Keras and the segmentation models library. The ResNet architecture was introduced to address the challenge of training intense neural networks by utilizing residual blocks. A residual block contains skip connections that allow the model to learn residual functions, making it easier to train deep networks. ResNet50, with its 50 layers, has a more profound architecture than other ResNet variants (ResNet18 or ResNet34), and it has demonstrated superior performance on various computer vision tasks, particularly in image classification and feature extraction. For this study, we used ResNet50 as the convolutional neural network architecture for getting tumor segments from brain mpMRI images. The pre-built model trained on the BraTS2020 dataset, which was downloaded from Kaggle and used to generate segmented outputs for comparative analysis. The pre-trained model is compiled with categorical cross-entropy loss, the Adam optimizer, and metrics such as accuracy and dice coefficient. A standalone ResNet50 model is also loaded with pre-trained weights and integrated as an encoder within the U-Net. A CSV logger is initialized for recording training metrics, and two callbacks, ReduceLROnPlateau and CSV logger, are employed for dynamic learning rate adjustments and logging training information. The final model summary provides a concise overview of the entire segmentation architecture, offering insights into the U-Net and the integrated ResNet50 encoder (Asiri et al., 2023; Khodadadi Shoushtari et al., 2022; Thhntb. (2021), n.d.).

### 2.3 Radiomics Feature Extraction

A total of 1213 radiomics features were extracted from all four modalities T1, T1ce, T2, and FLAIR. This resulted in a total of 4852 features extracted for every subject. The feature set of each modality for every subject includes 17 shapes, 18 first orders, 74 textures, 368 LoG (Laplacian of Gaussian), and 736 wavelet features. All feature calculations were performed using an open-source Python package called “*Pyradiomics*” version-3.0.1 (Van Griethuysen et al., 2017). The feature values were further z-normalized for statistical analysis. A detailed feature list is provided in **Table 1** and **Supplementary Table 1**.

**Table 1:** Features extracted using pyRadiomics.

|  |  |  |
| --- | --- | --- |
| **Shape** | VoxelVolume , MeshVolume , SurfaceArea , SurfaceVolumeRatio , Compactness1 , Compactness2 , Sphericity , SphericalDisproportion , Maximum3DDiameter , Maximum2DDiameterSlice , Maximum2DDiameterColumn , Maximum2DDiameterRow , MajorAxisLength , MinorAxisLength , LeastAxisLength , Elongation , Flatness | 17 |
| **First Order** | 10Percentile, 90Percentile, Energy, Entropy, InterquartileRange, Kurtosis, Maximum, Mean, Median, MeanAbsoluteDeviationMinimum, Range, RobustMeanAbsoluteDeviation, RootMeanSquared, Skewness, TotalEnergy, Uniformity, Variance | 18 |
| **Texture** | GLCM, GLRLM, GLSZM, GLDM, NGTDM | 74 |
| **LoG** | Sigma {2,3,4,5} | 368 |
| **Wavelet** | LLL, LLH, LHL, LHH, HLL, HLH, HHL, HHH | 736 |
| **Total Features (*per subject*)** |  | **1213** |

### 2.4 Performance Analysis

The performance analysis involved a dual approach. Initially, Dice Similarity Coefficient (DSC) and Hausdorff Distance (HD) were calculated using Python3 (Van Rossum & Drake, 2009), comparing the direct tumor segments obtained from CaPTk, 2DVNet, EnsembleUNets, and ResNet50 against a reference method. These metrics offered a granular assessment of the methods’ predictive accuracy, precision, and overall efficacy in segmenting the correspondence between predicted and reference segments. The Dice Similarity Coefficient (DSC) was calculated using the formula:

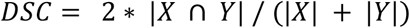

X and Y are reference, and the predicted segments sets, and when enclosed with vertical bars (e.g., |X|), it indicates the size, representing the number of elements in the respective set. The symbol ∩ denotes the intersection of two sets, signifying the common elements shared between them. Hausdorff Distance was calculated using :

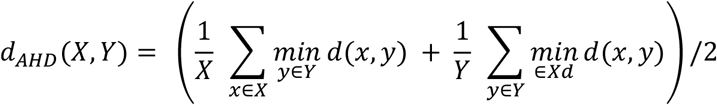

Here, the sets X and Y correspond to the voxels of the reference segments and the predicted segmentation, respectively (Aydin et al., 2021).

Subsequently, comprehensive statistical measures, including Concordance Correlation Coefficient (CCC), Total Deviation Index (TDI), Correlation, Root Mean Square Error (RMSE), and Coefficient of determination (R^2^), were computed using“DescTool”and“MethComp”packages in R software v4.2.1 (RStudio Team, 2020). Formulas used in above mentioned measures are given below:

1. CCC

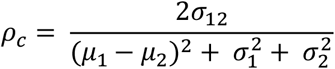

Agreement between reference method (variable 1) and other methods (variable 2) are characterized by Gaussian statistics. These variables have means μ1 and μ2, and standard deviations σ1 and σ2, with a covariance denoted as σ12.

2. R^2^

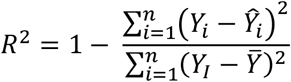

Here, *Y*_*i*_ is the reference value, *Ŷ*_*i*_ is the predicted value and 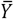 is the mean of the observed values.

3. RMSE

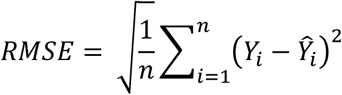

Here, *Y*_*i*_ is the reference value, *Ŷ*_*i*_ is the predicted value and n is the number of data points.

4. TDI

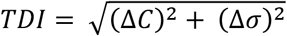

Here, ΔC is the difference in means, and Δ*σ* is the difference in standard deviations.

This multifaceted analysis aimed to compare the performance of the four reference segmentation methods across diverse dimensions, extracting features from individual subjects segmented by distinct methods. The results from both sets of analyses collectively provide valuable insights into which approach aligns most closely with the reference method, ensuring a robust selection for segmentation to yield reliable and accurate results. Additionally, feature wise analysis has also been done using R-packages that will provide a thorough understanding of each method’s performance on different feature category, aiding in the selection of the most reliable approach for brain tumor segmentation in diverse clinical scenarios. **Supplementary Table 2** contains the detailed statistical outcomes of these assessments.

## 3. Results

This study examines variations in the performance of methods for the automatic segmentation of tumor in brain mpMRI images. The study employs four different methods for segmenting brain tumors across 1251 subjects, each with four modalities. Each scan, consisting of 155 slices, involves labelling three sub-regions on each slice: Edema (ED), Necrosis/Non-enhancing tumor (NET), and Enhancing tumor (ET). Necrosis is identified in the hypointense region on T1ce, while interstitial fluid leakage indicative of edema is observed in hyperintense regions on FLAIR and T2. However, a few slices were selected to demonstrate the method’s performance graphically. In **Figure 3**, a single slice of T1ce and T2 MRI scans displays sub-region labels (necrosis outlined in red, enhancing tumor in blue/yellow, and edema in green) for both ground truth segments and tumor segments obtained from CaPTk, 2D-Vnet, EnsembleUNets, and ResNet50.

**Figure 3:**
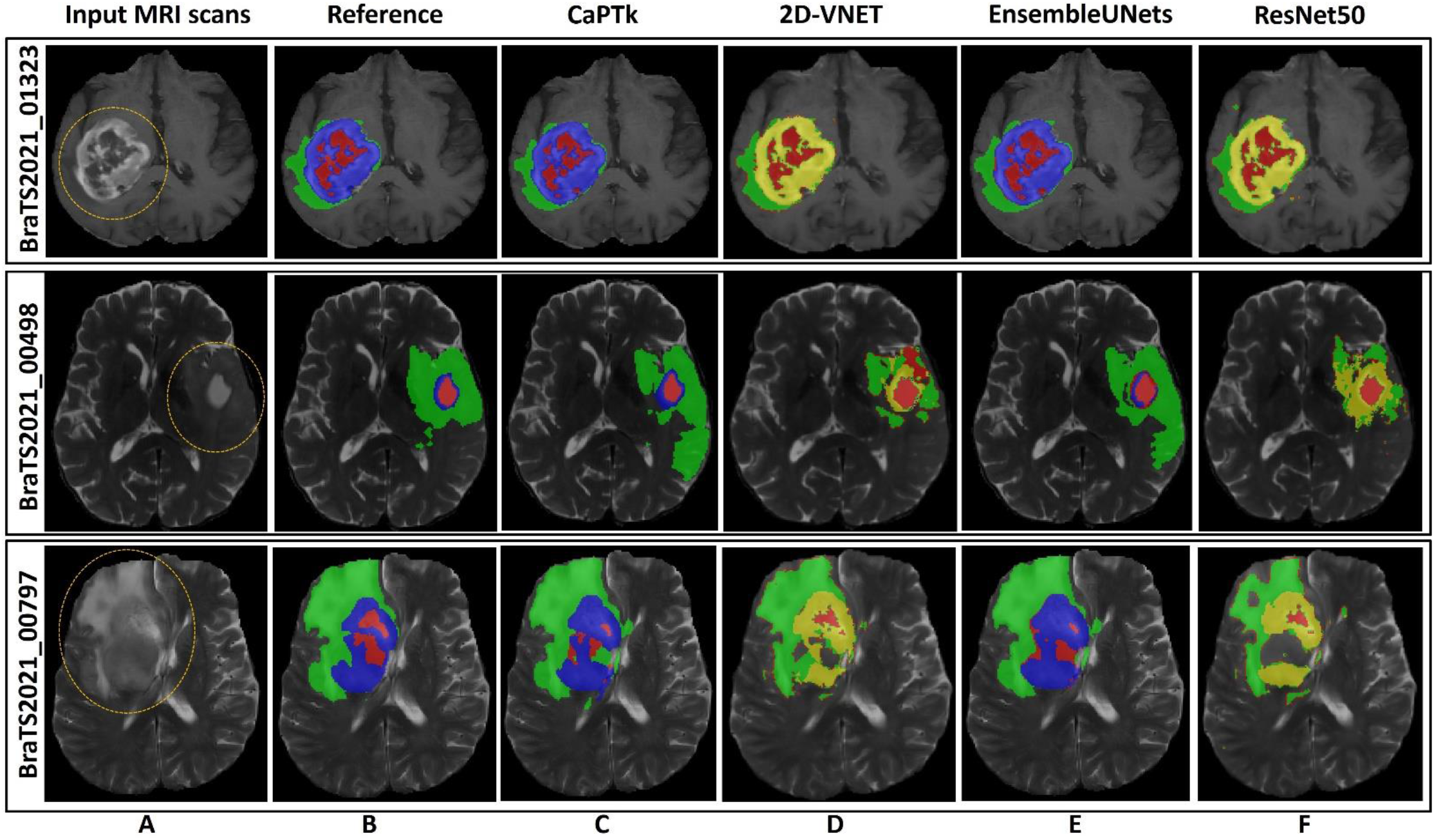
Segmentation Results Comparison. (A) Input MRI scans, (B) Reference tumor segment, and (C–F) segmentation outcomes by CaPTk, 2DVNET, EnsembleUNets, and ResNet50, respectively, for three representative samples from BraTS20212 dataset. This figure visually contrasts the segmentation performance of different methods against the reference, providing insights into their efficacy.

### 3.1 Performance comparison with state-of-the-art methods

#### 3.1.1 Performance-based on tumor segments

The assessment of segmentation methods, gauged by the Dice Similarity Coefficient (DSC) and Hausdorff Distance (HD), serves as a powerful indicator of their efficacy in delineating tumor boundaries (Kaur et al., 2023; Pemberton et al., 2023; Peng & Sun, 2023). A higher DSC signifies superior spatial overlap, indicating precise segmentation accuracy, while a lower HD reflects reduced spatial dissimilarity. EnsembleUNets demonstrated exceptional performance with a notable DSC of 0.93 and a minimal HD of 18.14, outperforming CaPTk (DSC: 0.91, HD: 32.95), 2DVNet (DSC: 0.006, HD: 105.2), and ResNet50 (DSC: 0.006, HD: 110) (**Table 2 and Supplementary Table 3**). These metrics collectively underscore how EnsembleUNets excels in capturing spatial congruence and minimizing spatial divergence, crucial for accurate and effective delineation of tumor subregions. Moreover, **Figure 4** presents graphical representations of the segmentation results, visually illustrating the comparative performance of the assessed method and emphasizing the EnsembleUNets’ superiority in tumor segmentation.

**Table 2:** Performance of four methods estimating precise segmentation compared with reference using segments. Values are presented as the average ± standard deviation (SD).

| Parameters | Dice Similarity Coefficient scores (DSC) | Hausdorff Distance (HD) |
| --- | --- | --- |
| **CaPTk** | 0.91 ± 0.11 | 32.95 ± 26 |
| **2DVNet** | 0.006 ± 0.02 | 105.2 ± 20 |
| **EnsembleUNets** | **0.93 ± 0.08** | **18.14 ± 18** |
| **ResNet50** | 0.006 ± 0.02 | 110 ± 19 |

**Figure 4:**
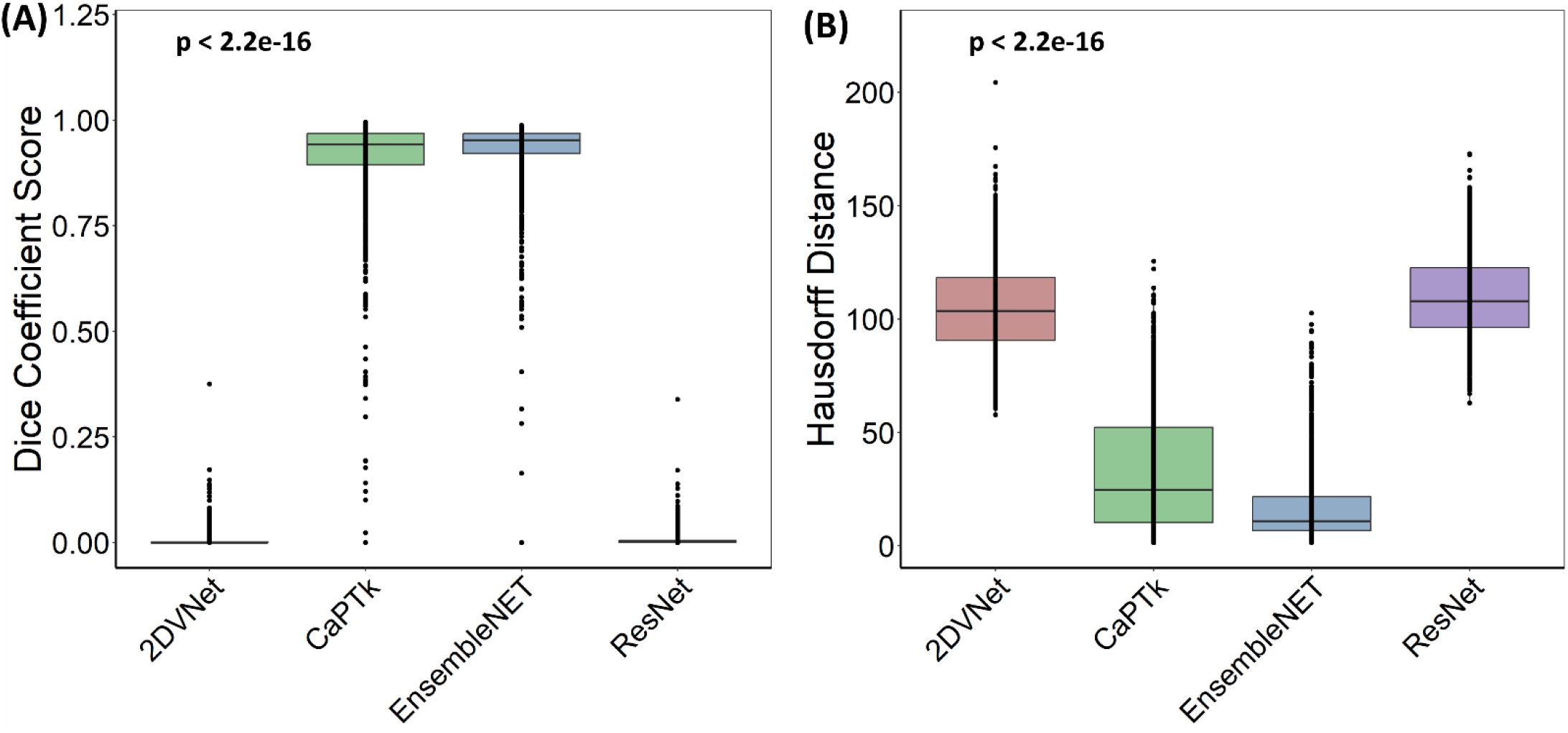
Boxplots of Dice Similarity Coefficient (DSC) scores and Hausdorff Distance (HD) metrics, illustrating the comparative performance of segmentation methods. The whiskers represent the range of scores, while the boxes denote interquartile ranges, providing a comprehensive view of segmentation accuracy across different methods.

#### 3.1.2 Performance-based on radiomics features

**Table 3** summarizes a comprehensive assessment of four segmentation method, wherein we examined a range of parameters to assess the performance of CaPTk, Vnet, EnsembleUNets, and ResNet. The distance metric, indicative of dissimilarity from the reference method, revealed EnsembleUNets as the frontrunner with the lowest distance value of 39.87, outclassing other methods, CaPTk (44.85), Vnet (69.136), and ResNet (65.523). EnsembleUNets achieved Concordance Correlation Coefficient (CCC) and correlation coefficients of 0.79, emphasizing its superior agreement with the reference data as compared to CaPTk (0.74), Vnet (0.452), and ResNet (0.50). EnsembleUNets also demonstrated a commendable coefficient of determination (R^2^: 0.67), surpassing CaPTk (0.60), Vnet (0.28), and ResNet (0.32). Furthermore, EnsembleUNets achieved lowest Root Mean Square Error (RMSE: 0.53), lowest among four methods. Total Deviation Index (TDI: 1.14) highlighted its closeness to the reference model compared to CaPTk (1.29), Vnet (1.98), and ResNet (1.88). Additionally, EnsembleUNets excelled in both Frobenius Norm (1561.85) and Spectral Norm (726.50), further underscoring its overall superior performance across a range of critical parameters in comparison to the other methods. These compelling findings underscore EnsembleUNets efficacy and accuracy, positioning it as a promising method for diverse medical imaging applications. To investigate the performance of the segmentation method further, we conducted a detailed assessment within five categories of radiomics features: Shape, First Order, Texture, Laplacian of gaussian (LoG), and Wavelet. Results of this analysis underscore EnsembleUNets dominance among the assessed methods CaPTk, 2DVNet, and ResNet50 in all five categories of radiomic features. EnsembleUNets consistently records the lowest distance metrices indicating its capability to closely align with the reference method in all categories of radiomics feature. Notably, EnsembleUNets excels in first-order features with the lowest distance (34.12), highest CCC (0.84), and superior predictive accuracy (R^2^: 0.74). The trend continues in texture, LoG, and wavelet features, where EnsembleUNets consistently demonstrates the most favourable metrics, including lower distances, higher CCC values, and superior accuracy measures (**Figure 5 and Supplementary Table 4**).

**Table 3:** Performance of four methods estimating precise segmentation compared with reference using radiomics features. Values in bold indicate best performance.

| Parameters* | CaPTk | 2DVNet | EnsembleUNets | ResNet50 |
| --- | --- | --- | --- | --- |
| Distance | 44.85 | 69.136 | **39.87** | 65.523 |
| CCC | 0.74 | 0.452 | **0.79** | 0.50 |
| Correlation | 0.74 | 0.452 | **0.79** | 0.50 |
| R<sup>2</sup> | 0.60 | 0.28 | **0.67** | 0.32 |
| RMSE | 0.59 | 0.84 | **0.53** | 0.80 |
| TDI | 1.29 | 1.98 | **1.14** | 1.88 |
| Frobenius Norm | 1717.10 | 2493.46 | **1561.85** | 2378.04 |
| Spectral Norm | 811.24 | 1104.60 | **726.50** | 890.64 |

\*CCC: Concordance Correlation Coefficient; R<sup>2</sup>: Coefficient of determination; RMSE: Root Mean Square Error; TDI: Total Deviation Index

**Figure 5:**
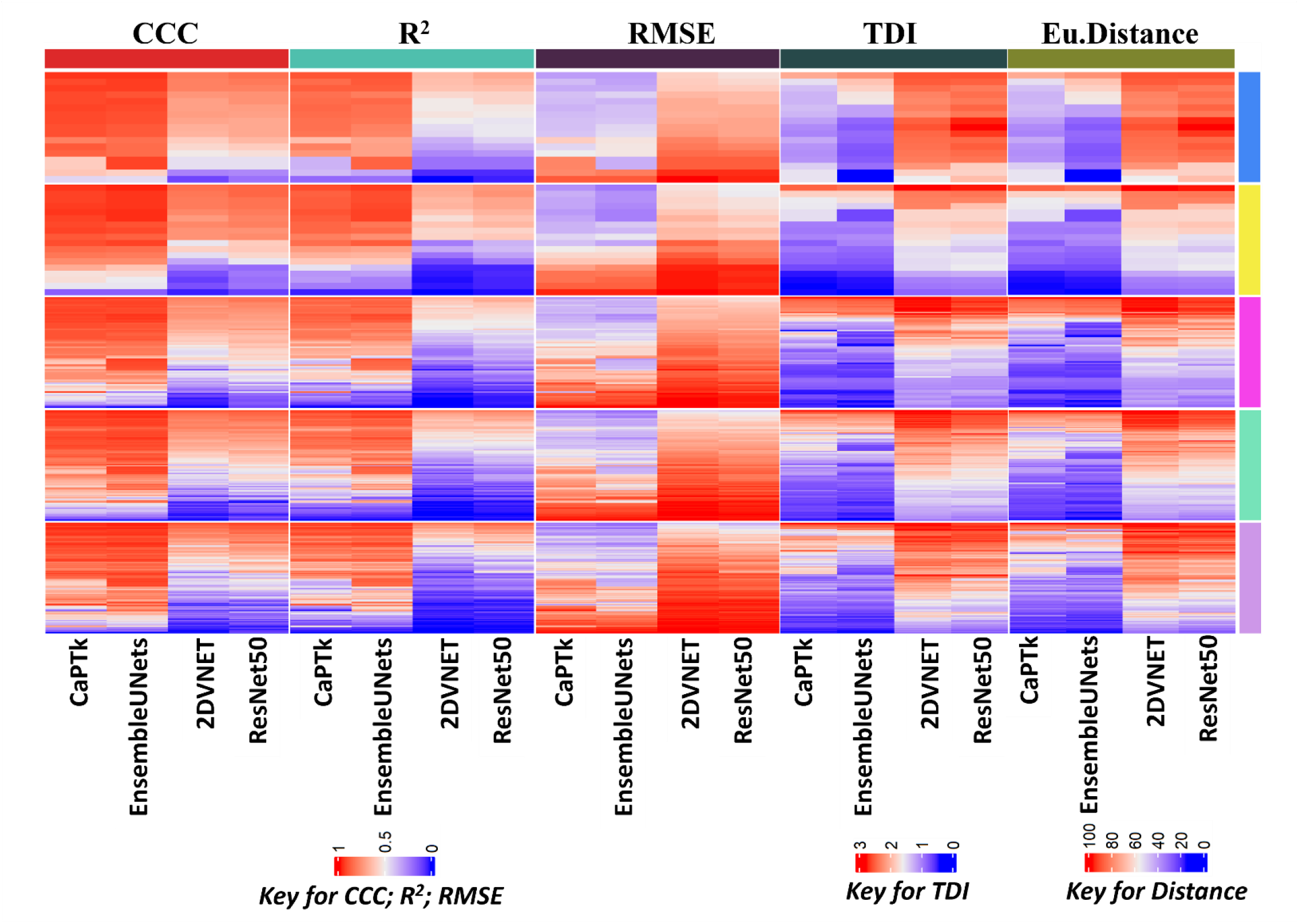
Comparative Performance of CaPTk, 2DVNet, EnsembleUNets, and ResNet50 across different radiomics feature categories. Top phenobar represents different performance metrics such as CCC (Concordance Correlation Coefficient), R^2^ (Coefficient of determination), RMSE (Root Mean Square Error), TDI (Total Deviation Index), and Euclidean Distance. Right phenobar corresponds to a specific feature category (blue for shape, yellow for first order, pink for texture, cyan for LoG, and plum for Wavelet) and indicates the method’s performance using a color-coded scale of different range.

## 4. Discussion

Multimodal MR imaging is now established as a fundamental technique in clinics. It is simple and yet sophisticated enough to detect brain tumor. MR imaging led to the origin of radiomics, where quantitative features are being extracted from the multimodal MR images. The accuracy of these features is contingent upon the precise segmentation of tumors from MR images. While numerous deep learning methods have been developed for brain tumor segmentation, each providing varying levels of accuracy, but none have been benchmarked yet (Kaur et al., 2023). In this study, we assessed the performance of leading and widely used brain tumor segmentation methods, aiming to benchmark the best among them by comparing them with the reference method. The core findings of this study can be summarized concisely through two points Firstly, we conducted segmentation on 1251 samples of BraTS2021 multimodal data employing the CaPTk, 2DVNet, EnsembleUNets, and ResNet50 methods. Secondly, we evaluated the performance of each method using diverse statistical metrics. Each sample in the BraTS2021 dataset consisted of four mpMRI images (T1, T2, T1ce, and FLAIR) and a pre-segmented image in nifti format, considered the reference. This reference method was derived from segmentation employing three DL-based methods and manual curation by a radiologist. Post-segmentation feature extraction from each modality using *Pyradiomics*, yielding 1213 features per modality per sample and 4852 features per subject. Various performance metrics were employed to determine which method produced results closest to the reference method by the dual approach (i.e., direct segments and radiomic features). The comprehensive evaluation consistently underscores EnsembleUNets as the preeminent method, showcasing superior performance across all metrics. Further, the feature-wise assessment again highlighted EnsembleUNets’ superior performance compared to CaPTk, 2DVNet, and ResNet50. This method exhibited exemplary performance across a spectrum of quantitative metrics, establishing its superiority as the preferred method of choice for brain tumor segmentation. It’s noteworthy that DeepMedic approach, operating at the CaPTk backend, demonstrated comparable performance to EnsembleUNets.

The plausible reason for the superior performance of EnsembleUNets is its unique combination of three methods (3D U-Net, 3D MI-U-Net, and 3D+2D MI-U-Net), leveraging different modalities and effective post-processing that provides a comprehensive and versatile approach. Initially, the 3D MI-UNet is a trained for predicting the training data, followed by employing the 2D slices from four multi-modal MR images, BP, and probability maps of WT, TC, and ET as inputs for training a subsequent 2D MI-UNet. This sequential training strategy optimally combines the strengths of both 3D and 2D representations and results in improved accuracy and robustness.

In the realm of medical diagnosis, our benchmarked method, EnsembleUNets, provides a reliable foundation for identifying and characterizing tumor subregions. The robust segmentation it achieves not only opens a window for a more intricate interpretation of the disease but also unveils an opportunity to employ this benchmarked method for precise tumor classification and grading. Moreover, in the context of treatment planning, the superior performance of EnsembleUNets ensures an accurate delineation of tumor boundaries. This precision is particularly crucial in radiation therapy, where optimal treatment plans rely on precise tumor localization. EnsembleUNets based segmentation might contributes to more targeted and individualized treatment strategies, maximizing therapeutic efficacy while minimizing potential harm to surrounding healthy tissues.

Moreover, integration of EnsembleUNets as the preferred choice for brain tumor segmentation can establish a solid foundation for future explorations of relationship between radiomic and genomic features. This method, exhibiting exemplary performance across a spectrum of quantitative metrics, paves the way for advancements in better disease management and treatment planning.

## 5. Conclusion

In this study, we have benchmarked the four widely used brain tumor segmentation methods. As imaging hold prominent role in diagnosis and treatment planning of glioma, this benchmarking study will be critical to choose the right segmentation method. To best of our knowledge, this is the first study of its kind which used comprehensive radiomic features extracted from the tumor segments for the comparison analysis of tumor segmentation methods. The findings of this study demonstrated that EnsembleUNets outperformed other segmentation methods employed, showcasing its higher segmentation precision. This method stands out as a most accurate, robust, and user-friendly method for the researchers and clinicians, enabling accurate tumor segmentation and offering valuable insights for informed treatment strategies. Based on our findings, we recommend to use EnsembleUNets for the segmentation of brain tumor from multimodal MR images.

## Data Availability

The dataset analysed in this study is available in BraTS2021 dataset: http://braintumorsegmentation.org/

## 6. Data Availability

The dataset analysed in this study is available in BraTS2021 dataset: *http://braintumorsegmentation.org/*.

## 7. Author Contribution

RK and KK conceived the idea. KK performed all the computational and statistical analysis in the supervision of RK. KVR assisted in understanding of MR images. AM assisted the statistical analysis. NK assisted in segmentation of MR images. KK and RK wrote the manuscript.

## 8. Acknowledgments

Authors acknowledge the infrastructure provided by the Indian Institute of Technology Hyderabad. KK acknowledge the financial support from the Ministry of Education (MoE), India.

